# Mechanical Thrombectomy in Extended Time Window: Real-World Data from a Colombian Stroke Center

**DOI:** 10.1101/2025.05.05.25327039

**Authors:** Andrés Y. Vásquez, Camilo S. Alvarado-Bedoya, María F. Estévez-Ochoa, Adriana Reyes-Gonzalez, Melquizidel Galvis, Carlos A. Ferreira, Oliverio Vargas, Daniel Mantilla

**Affiliations:** Department of Interventional Radiology, Universidad Autónoma de Bucaramanga, Bucaramanga, Santander, Colombia; Department of Interventional Radiology, Fundación Oftalmológica de Santander-Clínica Ardila Lülle, Floridablanca, Santander, Colombia; Clinical Research Group-UNAB, Universidad Autónoma de Bucaramanga, Bucaramanga, Colombia

**Keywords:** Endovascular thrombectomy, Ischemic stroke, Extended window stroke, Perfusion imaging, Neurointervention, Colombia

## Abstract

**Background:** Mechanical thrombectomy (MT) has revolutionized the management of acute ischemic stroke (AIS) due to large vessel occlusion (LVO), particularly when performed within the first 6 hours after symptom onset. The development of perfusion imaging software has enabled patient selection to thrombectomy for up to 24 hours in selected cases with salvageable brain tissue, following the criteria of trials such as DEFUSE 3. However, the real-world application of these criteria, remains understudied.

**Objective:** To determine the clinical outcomes of patients with AIS treated with MT in an extended window (>6 hours), comparing patients who met DEFUSE 3 perfusion criteria versus those who did not.

**Methods:** A retrospective analysis was conducted on patients undergoing MT between 6 and 24 hours after symptom onset. Clinical outcomes were assessed at hospital discharge and 90 days using the modified Rankin Scale (mRS). Patients were divided into two groups based on whether they met DEFUSE 3 perfusion criteria.

**Results:** A total of 80 patients were treated. Median age was 76 years (IQR 62,5-83). Wake–up strokes accounted for 45. Median ASPECTS score was 7 (SD 2.25) and median ischemic core volume was 18.9 ml (IQR 8,2 – 44,7). Of 76 patients analyzed, 37 (48.7%) met DEFUSE 3 criteria and 39 (51.3%) did not. Although patients meeting the criteria showed a trend toward better functional outcomes (mRS 0–3 at discharge: 66.6% vs 33.3%; p = 0.14), similar outcomes were observed at 90 days (63.6% vs 36.3% p = 0.34). Additionally, the group that did not meet DEFUSE 3 criteria had a higher proportion of wake-up strokes (p = 0.02), a relevant factor in extended-window decision-making. No statistically significant differences were found in mortality or severe disability between groups.

**Conclusion:** In our study a good functional outcomes was more frequent in patients selected by DEFUSE 3 perfusion criteria, but a notable number of patients outside these criteria also achieved functional recovery. These findings support a more flexible and context-aware approach to patient selection in extended windows. Future prospective studies should aim to refine patient selection protocols that balance safety, efficacy, and accessibility.

## Introduction

Stroke is the second leading cause of death globally, accounting for nearly 7 million deaths worldwide. It is also the third leading cause of disability with its incidence rising due to population growth and aging^1^, Approximately 80% of strokes affect the anterior circulation, while 20% involve posterior circulation^2^.

In 2023, the Integrated Social Protection Information System (SISPRO), recorded 118,438 cases of cerebrovascular accidents in Colombia, with 87.9% being ischemic strokes and 12.1% to hemorrhagic strokes. Stroke-related mortality in Colombia during the same period, according to the National Department of Statistics, was 16,946 patients, representing 16.2% of the total national mortality, making it the second leading cause of non-violent death in the country.

Effective stroke management demands a multidisciplinary approach and prompt imaging studies in the emergency department to enable a rapid and effective approach^3^, enabling the decision to provide a pharmacological and/or endovascular treatment alternative.

In 2015 studies such as MR CLEAN^4^, SWIFT PRIME^5^, REVASCAT^6^, ESCAPE^7, and^ EXTEND IA^8^ demonstrated that endovascular thrombectomy reduces disability in ischemic strokes due to acute large vessel occlusion. The HERMES^8^ meta-analysis later confirmed these findings, reporting a number needed to treat (NNT) of 2.6 within the 6-12 hours^1,9^.

However, many patients were ineligible for endovascular treatment until the 2018 DEFUSE 3^10^ and DAWN^11^ trials, which demonstrated favorable outcomes in select patients assessed via perfusion computed tomography or magnetic resonance imaging. These studies reported an NNT of 2 and 2.8, respectively, extending the treatment window to 24 hours.

The inclusion criteria guided by perfusion in the DEFUSE 3 study were in patients with carotid or M1 segment middle cerebral artery occlusion, NIHSS > 6, a Rankin score <2, and a Target mismatch profile on CT or MR perfusion imaging, as determined by an automated image post-processing system: with infarct core volume <70 ml, mismatch volume >15 ml (Tmax >6 s), and mismatch ratio (penumbra/core) >1.8^10^.

Although current studies do not fully encompass all patients with late-window stroke who undergo thrombectomy and their associated outcomes, further investigation is warranted to address this gap.

This retrospective cohort study analyzes the number of patients admitted with late-window stroke, identifies those who underwent mechanical thrombectomy, evaluates their clinical outcomes, complications, and compare patients who met perfusion-guided criteria with those who did not, providing insight into the impact of these selection criteria.

## Materials and methods

### Data source and patient selection

An anonymized database from the Interventional Radiology Department of the Fundación Oftalmológica de Santander - FOSCAL and Fundación FOSUNAB will be used, from which the variables of interest collected between the years 2020 and 2024 will be extracted.

Patients included in the study underwent mechanical thrombectomy for ischemic stroke beyond the pharmacological window (6-24 hours). They were assessed using CT perfusion imaging and classified based on whether they fulfilled the imaging criteria of the DEFUSE 3 study or not, CT perfusions studies were performed using 16, 80 and 320 slice scanners, automatically processed using OLEA software.

Patients over 18 years old with ischemic cerebrovascular events affecting vascular structures of the anterior circulation (internal carotid artery, middle cerebral artery in the M1 or M2 segment, or anterior cerebral artery in the A1 or A2 segment) were included. The diagnosis was confirmed by CT angiography and CT perfusion imaging, with or without prior thrombolytic therapy, and with an evolution of the ischemic event within up to 24 hours from symptom onset.

Exclusion criteria included intracranial bleeding, hemorrhagic transformation of the infarcted area, or posterior circulation involvement.

A detailed calculation was conducted to determine the required sample size based on the DEFUSE3 study. A difference in proportions approach was used, with a significance level (alpha) set at 0.05 and a power of 0.80. The expected difference (delta) between proportions was defined as 43.1%, based on the rate of complete recanalization at 24 hours in the group of patients who received endovascular treatment. Based on these parameters, the estimated total sample size was calculated to be N = 70 participants.

### Data characteristics

The variables considered included age, time of evolution, NIHSS score at admission, whether it was a wake-up stroke, prior thrombolysis administration, ASPECTS score on non-contrast CT, occlusion site according to CT angiography or diagnostic angiography, infarct volume on perfusion imaging, mismatch between the penumbra core/ischemic core in the perfusion study, post-thrombectomy revascularization score (TICI), functional outcome, and survival.

### Statistical analysis

For the univariate analysis, qualitative variables will be described using absolute and relative frequencies (percentages) along with their respective confidence intervals. Quantitative variables will be presented with measures of central tendency and dispersion, such as mean and standard deviation, or median and interquartile range, depending on the data distribution (the distribution will be assessed using the Shapiro-Francia test).

## Results

### Baseline characteristics and comorbidities

A total of 80 patients were included (table 1). The median age was 76 years (IQR 62.5–83), with a predominance of males (56.25%, 95% CI: 45.01–66.87). Hypertension (HTN) was the most prevalent risk factor, affecting 72.5% of the patients (95% CI: 61.48–81.31), followed by atrial fibrillation (21.2%, 95% CI: 13.50–31.81) and diabetes mellitus (17.5%, 95% CI: 10.52–27.66). The median time since symptom onset was 11.5 hours (IQR 7–13.9). Mean NIHSS at admission was 14 (SD 6.69), indicating moderate-to-severe neurological impairment. Wake-up strokes accounted for 45% (95% CI: 34.28–56.20) of cases, with a baseline mRS score of 0 (SD 0.76).

**Table 1.** Baseline Characteristics.

|  |  |
| --- | --- |
| Baseline characteristics |  |
| Median age | 76 (IQR 62,5-83) |
| Risk factors<br>Hypertension<br>Diabetes Mellitus<br>Atrial fibrillation<br>Ischemic cardiomyopathy<br>Previous ischemic stroke | 58 - 72,5% (95% CI 61,48 – 81,31)<br>14 - 17,5% (95% CI 10,52 – 27,66)<br>17 - 21,2% (95% CI 13,50 – 31,81)<br>5 - 6,25% (95% CI 2,57 – 14,39)<br>10 – 12,5% (95% CI 6,76 – 21,94) |
| Time from stroke onset | 11.5 (IQR 7 – 13,9) |
| NIHSS | 14,5 (SD 6,69) |
| Wake – up stroke | 36 – 45% (95% CI 34,28 – 56,20) |
| Baseline Rankin score | 0 - (SD 0,76) |
| ASPECTS | 7,28 (SD 2,25) |
| Perfusion variables<br>Ratio<br>Ischemic core -- flow <25% of contralateral<br>Penumbra - TTP > 5<br>Difference core/penumbra<br>Relative mismatch | 2,34 (IQR 1,5 – 4,75)<br>18,9 (IQR 8,2 – 44,7)<br>58,8 (IQR 28,4 – 95,4)<br>30,2 (IQR 13,8 – 51,1)<br>58,1 (IQR 34,9 – 79,1) |
| Large vessel occlusion (LVO) | 60 – 75% (95% IC 64,13 – 83,42) |
| Oclusionion LOCATION<br>TANDEM<br>Internal carotid<br>M1<br>Medium vessel | 24 – 30% (95% CI 20,81 – 41,12)<br>8 – 10% (95% CI 5,00 – 18,98)<br>28 – 35% (95% CI 25,18 – 46,26)<br>20 – 25% (95% CI 16,57 – 35,86) |
| TREATMENT CHARACTERISTICS |  |
| Door-to-groin time | 80 min (IQR 67–122.5) |
| Thrombectomy<br>Aspiration<br>Aspiration + stent retriever | 30 – 37,5% (95% CI 27,42 – 48,79)<br>39 – 48,75% (95% CI 37,80 – 59,82) |
| Number of passes (mean) | 2.57 (SD 1.23) |
| TICI<br>0 | 3 – 3,75% (95% IC 1,18 – 11,24)<br>2 – 2,5% (95% IC 0,60 – 9,71) |
| • 1<br>• 2 a<br>• 2 b<br>• 2 c<br>• 3 | • 10 – 12,5% (95% IC 6,76 – 21,94)<br>• 20 – 25% (95% IC 16,57 – 35,86)<br>• 4 – 5% (95% IC 1,84 – 12,82)<br>• 41 – 51,2% (95% IC 40,17 – 62,19) |
| ICU Length of stay | 5 (IQR 3 – 7,5) |
| Hospital Length of stay | 9,5 (IQR 6 – 18,5) |
| TOAST<br>• Atherosclerosis<br>• Cardioembolic<br>• Lacunar<br>• Determined<br>• Undetermined | • 7 – 9,09 % (95% IC 4,32 – 18,12)<br>• 38 – 49,35% (95% IC 38,15 – 60,61)<br>• 1 – 1,3% (95% IC 0,17 – 9,00)<br>• 7 - 9,09 % (95% IC 4,32 – 18,12)<br>• 24 - 31,17% (95% IC 21,66 – 42,58) |

Imaging findings provided key insights into the diagnostic evaluation of extended-window stroke patients. The mean ASPECTS score was 7.28 (SD 2.25), suggesting a moderate extent of early ischemic changes on non-contrast CT imaging.

Perfusion imaging revealed the following key parameters: Mean perfusion ratio: 2.34 (IQR 1.5–4.75), median ischemic core volume: 18.9 mL (IQR 8.2–44.7), median penumbra volume: 58.8 mL (IQR 28.4–95.4), median core-penumbra difference: 30.2 mL (IQR 13.8–51.1). Relative mismatch: 58.1% (IQR 34.9–79.1).

These findings indicate substantial salvageable brain tissue (penumbra) in most cases, supporting the feasibility of intervention in extended time windows. Large vessel occlusion (LVO) in carotid or M1 segments, was identified in 75% of cases (95% CI: 64.13–83.42).

Tandem occlusions accounted for 30% (95% CI: 20.81–41.12). M1 occlusions were the most frequent, representing 35% (95% CI: 25.18–46.26). Distal occlusions occurred in 25% (95% CI: 16.57–35.86).

The median door-to-groin time was 80 minutes (IQR 67–122.5). The mean number of passes required for thrombectomy was 2.57 (SD 1.23).

Aspiration was used in 37.5% (95% CI: 27.42–48.79). Combined aspiration and stent retriever techniques were more common, utilized in 48.75% (95% CI: 37.80– 59.82). Recanalization with a TICI score between 2b – 3 was made in more than 80% of patients.

The median stay in the ICU was 5 days (IQR: 3–7.5), while the median total hospital stay was 9.5 days (IQR: 6–18.5).

Etiological classification using the TOAST criteria identified cardioembolic stroke in 49.35% (95% CI 38.15-60.61). At discharge, 43.42% had moderate-to-severe disability (mRS 4-5) and 28.95% died (mRS 6). At 3-month follow-up (n=60), functional independence (mRS 0-2) was achieved in 30% of cases, while mortality remained high at 40%.

### Outcomes

Of the 80 patients treated, only 76 were included in the initial outcome analysis. The remaining 4 were transferred to another hospital after undergoing thrombectomy, preventing further follow-up. Among the 76 patients analyzed, 37 met the perfusion criteria while 39 did not. A 3-month follow-up was conducted on 60 patients, 31 from the perfusion criteria group and 29 from the non-criteria group. The outcomes were statistically similar both during hospitalization and at the 3-month follow-up.

When comparing patients (table 2 and 3) with and without a favorable perfusion profile (Defuse positive vs. Defuse negative), no significant differences were found in age (p = 0.84), time from symptom onset to treatment (median 11.8 h vs. 10.9 h), presence of large vessel occlusion (80.4% vs. 69.2%, p = 0.30), or administration of intravenous thrombolysis (p = 1.00). Likewise, the type of anesthesia used, the need for decompressive craniectomy, and door-to-puncture times were similar between groups.

**Table 2.**
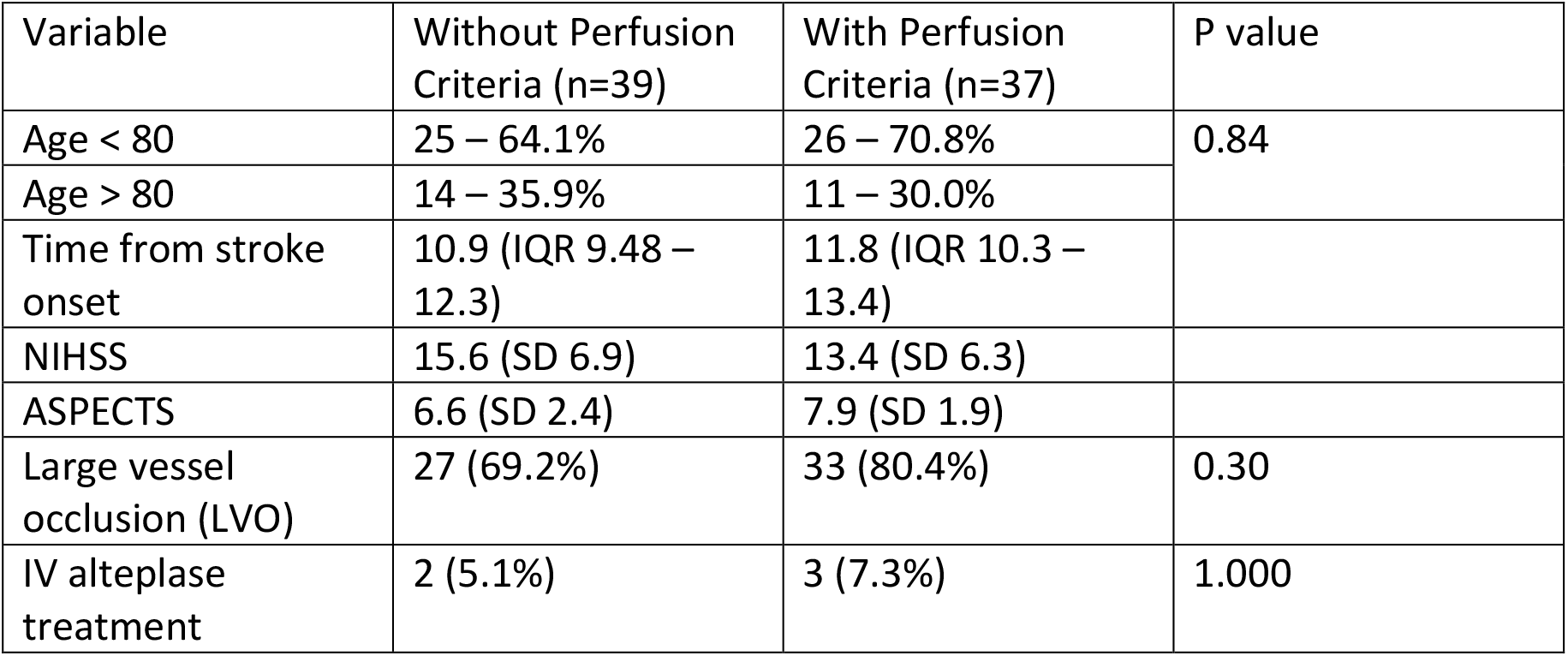
Groups baseline characteristics.

| Variable | Without Perfusion Criteria (n=39) | With Perfusion Criteria (n=37) | P value |
| --- | --- | --- | --- |
| Age < 80 | 25 – 64.1% | 26 – 70.8% | 0.84 |
| Age > 80 | 14 – 35.9% | 11 – 30.0% |  |
| Time from stroke onset | 10.9 (IQR 9.48 – 12.3) | 11.8 (IQR 10.3 – 13.4) |  |
| NIHSS | 15.6 (SD 6.9) | 13.4 (SD 6.3) |  |
| ASPECTS | 6.6 (SD 2.4) | 7.9 (SD 1.9) |  |
| Large vessel occlusion (LVO) | 27 (69.2%) | 33 (80.4%) | 0.30 |
| IV alteplase treatment | 2 (5.1%) | 3 (7.3%) | 1.000 |

**Table 3.** Procedural and Imaging Variables.

|  | Without Perfusion Criteria | With Perfusion Criteria | P value |
| --- | --- | --- | --- |
| Perfusion – Core/penumbra ratio | 2.70 (IQR 1.26–4.15) | 14.7 (IQR 2.5–26.9) |  |
| Ischemic core – rCBF <25% | 43.1 (IQR 30.8–55.3) | 19.2 (IQR 13.9–24.5) |  |
| Penumbra – TTP > 5 seg | 67.0 (IQR 45.9–88.0) | 68.7 (IQR 57.8–79.6) |  |
| Core/penumbra difference ml | 24.0 (IQR 10.9–37.2) | 49.5 (IQR 41.2–57.8) |  |
| Relative mismatch | 40.5 (IQR 33.3–47.7) | 73.4 (IQR 68.5–78.3) |  |
| Door-to-puncture time (min) | 103.1 (IQR 72.3–133.9) | 107.9 (IQR 88.0–127.8) |  |
| Anesthesia – Local | 1 – 2.5% | 1 – 2.4% | 0.81 |
| Anesthesia – Local + sedation | 22 – 56.4% | 26 – 63.4% |  |
| Anesthesia – General | 16 – 41.0% | 14 – 34.1% |  |
| Number of passes (mean) | 2.9 (SD 1.39) | 2.2 (SD 1.0) |  |
| Passes – 1 | 8 – 25.0% | 10 – 26.3% | 0.09 |
| Passes – 2 | 3 – 9.3% | 11 – 28.9% |  |
| Passes – ≥3 | 21 – 65.6% | 17 – 44.7% |  |
| Intraarterial thrombolysis | 8 – 20.5% | 4 – 9.7% | 0.22 |
| TICI ≥ 2b | 31 – 79,3% | 34 – 82,8 | 0,81 |

Patients with a favorable perfusion profile showed slightly lower baseline NIHSS scores (13.4 vs. 15.6) and higher ASPECTS scores (7.9 vs. 6.6), suggesting a less severe initial clinical and radiological presentation. Notably, perfusion imaging revealed a significantly smaller core infarct volume (median 19.2 ml vs. 43.1 ml) and a larger mismatch volume (49.5 ml vs. 24.0 ml) in the Defuse positive group, along with a greater relative mismatch ratio (73.4% vs. 40.5%). These findings support the hypothesis that patients with more salvageable tissue may benefit more from mechanical thrombectomy, even in extended time windows.

Procedurally, the Defuse positive group required fewer device passes on average (2.2 vs. 2.9) and achieved a slightly higher rate of successful recanalization (TICI ≥ 2b: 82.8% vs. 79.3%), although these differences did not reach statistical significance (p = 0.81). This may reflect more favorable vascular anatomy or less thrombus burden in patients with a positive perfusion profile.

The multivariate analysis revealed relevant trends in functional outcomes and mortality according to key clinical and radiological variables.

Hospital discharge mRS scores (table 4) also trended in favor of perfusión criteria patients (functional: 37.8% vs. 17.9%, *p* = 0.14) and the difference continue at 90 days, the limited sample size may have precluded statistical significance. This supports the hypothesis that favorable perfusion imaging profiles may help identify patients who are more likely to benefit from reperfusion therapies even beyond conventional time windows. Notably, patients without perfusión criteria that no present wake-up stroke demonstrated a significant association with unfavorable outcomes (*p* = 0.02). This suggests that this group have a high-risk presentation and poor outcomes. In contrast, large vessel occlusion (LVO) and TICI ≥ 2b reperfusion status did not demonstrate statistically significant associations with outcomes (*p* = 0.22 and *p* = 0.083, respectively), though trends suggest better functional outcomes in the presence of successful reperfusion and LVO.

**Table 4.** Multivariate analisis.

| Comparison | Without Perfusion Criteria | With Perfusion Criteria | P value |
| --- | --- | --- | --- |
| mRs at hospital discharge |  |  | 0.14 |
| Functional (0–3) | 7 – 17.9% | 14 – 37.8% |  |
| Moderate-Severe (4–5) | 20 – 51.2% | 13 – 35.1% |  |
| Death (6) | 12 – 30.7% | 10 – 27.0% |  |
| NO wake-up stroke |  |  | 0.02 |
| Functional (0–3) | 4 – 16.0% | 8 – 50.0% |  |
| Moderate-Severe (4–5) | 12 – 48.0% | 7 – 43.7% |  |
| Death (6) | 9 – 36.0% | 1 – 6.2% |  |
| Large vessel occlusion (LVO) |  |  | 0.22 |
| Functional (0–3) | 4 – 14.8% | 10 – 34.8% |  |
| Moderate-Severe (4–5) | 11 – 40.7% | 9 – 31.0% |  |
| Death (6) | 12 – 44.4% | 10 – 34.4% |  |
| TICI ≥ 2b |  |  | 0.083 |
| Functional (0–3) | 6 – 19.3% | 13 – 40.6% |  |
| Moderate-Severe (4–5) | 19 – 61.2% | 11 – 34.3% |  |
| Death (6) | 6 – 19.3% | 8 – 25.0% |  |
| mRs at 90 days |  |  | 0.34 |
| Functional (0–3) | 8 – 27.5% | 14 – 45.1% |  |
| Moderate-Severe<br>(4-5) | 8 – 27.5% | 6 – 19.3% |  |
| Death (6) | 13 – 44.8% | 11 – 35.4% |  |

## Discussion

This study analyzes the management of extended-window stroke patients at one of the few referral centers in northeastern Colombia. Access to treatment in this region is unequal, as it is only available in capital cities, posing significant challenges in education and logistics. These findings align with studies from Latin America, where limited human and financial resources may lead to an underestimation of the true impact of this condition^12,13^.

Compared to the DEFUSE 3 trial^10^, some differences were observed in our study: Treatment window: 6–16 hours vs. 6–24 hours. Our study included large and medium vessel occlusion. NIHSS score: 16 vs. 15 points. Wake-up strokes: 53% vs. 45%. Perfusion imaging: Ischemic core: 9.4 mL (IQR 2.3–25) vs. 18.9 mL (IQR 8.2–44.7), Penumbra volume: 114.7 mL (IQR 79.3–146) vs. 58.8 mL (IQR 28.4– 95.4). ASPECTS score: 8 vs. 7 points. Symptom onset to treatment: 10.5 vs. 11.5 hours. Door-to-groin time: 59 vs. 80 minutes.

At the 3-month follow-up Mortality was 14% in DEFUSE 3 vs. 35% (patients with perfusion criteria) and 44.3% (those who did not). Functional independence (mRS 0–2) was 45% in DEFUSE 3 vs. 29.9% in our study, but mRS 0–3 in our study in patients meeting perfusion criteria were 45% vs. 27.5% in those who did not. Intracranial hemorrhage (symptomatic or not): 16% vs. 27.3% in this study.

Additionally, we included variables not considered in DEFUSE 3, such as medium-vessel thrombectomy, which had an mRS 0–3 rate of nearly 50%, though statistical significance was not reached due to the small sample size but is a similar outcome than the STAR study (46,7%)^14^ . In patients requiring craniectomy (8 cases), 6 (75%) had a severe disability (mRS 4–5), and 2 died, those results concerning the evidence^15,16^.

Patients meeting perfusion criteria tended to have better outcomes; however, our findings reveal that a significant proportion of patients who did *not* meet DEFUSE 3 criteria also achieved functional Independence.

This observation is particularly relevant for institutions where access to advanced perfusion software or automated mismatch quantification is limited. In these settings, decisions are often based on a combination of clinical judgment, non-contrast CT, and CT angiography. The recovery seen in these patients raises questions about the rigidity of current criteria and underscores the importance of considering other factors such as collateral circulation, clinical presentation, and individual brain resilience.

Although our results did not reach statistical significance in several comparisons, likely due to sample size limitations, they suggest a clear trend. These findings contribute to a growing body of evidence supporting the use of MT beyond rigid selection rules, especially in real-world settings.

## Conclusions

In our study a good functional outcomes was more frequent in patients selected by DEFUSE 3 perfusion criteria, but a notable number of patients outside these criteria also achieved functional recovery. These findings support a more flexible and context-aware approach to patient selection in extended windows. Future prospective studies should aim to refine patient selection protocols that balance safety, efficacy, and accessibility.

## Data Availability

all the data is available, if something aditional is needed we can provide it.

## Tables – Perfusion Criteria Study

